# Mathematical modelling based study and prediction of COVID-19 epidemic dissemination under the impact of lockdown in India

**DOI:** 10.1101/2020.07.25.20161885

**Authors:** Vipin Tiwari, Namrata Deyal, Nandan S. Bisht

## Abstract

COVID-19 (SARS-CoV-2) is rapidly spreading in South Asian countries especially in India. India is the fourth most COVID-19 affected country at present (as on July 10, 2020). With limited medical facilities and high transmission rate, study of COVID-19 progression and its subsequence trajectory need to be analyzed in India. Epidemiologic mathematical models have the potential to predict the epidemic peak of COVID-19 under different scenario. Lockdown is one of the most effective mitigation policy adapted worldwide to control the transmission rate of COVID-19 cases. In this study, we use an improvised five compartment mathematical model i.e. Susceptible (S) - exposed (E)- infected (I)- recovered (R)- death (D) (SEIRD) to investigate the progression of COVID-19 and predict the epidemic peak under the impact of lockdown in India. The aim of this study to provide the most accurate prediction of epidemic peak and to evaluate the impact of lockdown on epidemic peak shift in India. For this purpose, we examine most recent data (up to July 10, 2020) to enhance the accuracy of outcomes from the proposed model. The obtained results indicate that COVID-19 epidemic peak would appear on around mid-August 2020 in India and corresponding estimated cases would be 2.5×10^6^ under current scenario. In addition, our study indicates that existence of under-reported cases (∼10^5^) during post-lockdown period in India. It is expected that nationwide lockdown would lead to epidemic peak suppression in India. The obtained results would be beneficial for determining further COVID-19 mitigation policies not only in India but globally.

## INTRODUCTION

COVID-19 is a contagious disease of SARS-COV family [1]. It is responsible for the biggest health crisis of 21^st^ century across the globe in 2020. COVID-19 infected patients generally exhibit common symptoms like cough, fever and respiratory disorders [2]. The most serious issue with COVID-19 is its drastic growth rate in world. Therefore, it has been declared as a global pandemic by World Health Organization (WHO) on March 11, 2020 just after two months since its first case reported in Wuhan (China) [3]. At present (as on July 10, 2020), total confirmed cases in world are 1.2×10^7^ and total number of deaths are 5.5×10^5^ [4]. United States of America (USA), Brazil, Russia, India and United Kingdom (UK) are the top five most affected countries as on July 10, 2020 [4]. Moreover, as the official vaccination of COVID-19 is not available till date, lockdown has been implemented as a preliminary containment policy in most of COVID-19 affected countries. World Health Organization (WHO) reported data indicates that lockdown is an effective policy for controlling COVID-19 spread [4]. Despite of several mitigation policies, COVID-19 cases are in alarming situation in south Asian countries particularly in India. Being a developing nation and second largest populated country, it is very challenging to control the spread of COVID-19 in India. The first COVID-19 confirmed case was found in Kerala on January 30, 2020 [6]. Under the lack of sufficient medical facilities in a country of 1.3 billion people, various containment strategies (international travel restrictions, nationwide lockdown etc.) have been implemented by government of India (March 25-May 31, 2020) in various phases [7, 8]. On June 01, 2020 onwards, the total lockdown has been partially lifted in India. It resulted as sudden growth in COVID-19 cases in last forty days. At present, COVID-19 cases are dramatically increasing in India and tending towards its epidemic peak [9]. Therefore, this particular time (June 2020) is very critical for estimating the subsequent trajectory of COVID-19 in India.

Mathematical models have the potential to trace and predict the epidemic trajectory under different scenario. Various mathematical, statistical models have been proposed to understand the dissemination trajectory for a pandemic [10-17]. Among these models, Susceptible(S)-Infected (I) - Recovered(R) model (SIR model) has been frequently used in past to predict the influence of HIV virus [10], plague [11], SARS [12] etc. Recently, SIR model has also been applied for prediction of COVID-19 trajectory and its epidemic peak [13-22]. However, such studies have been carried out at the very earlier stage of pandemic. Moreover, such studies are primarily focussed on the COVID-19 spread tracing under normal circumstances i.e. containment strategies (policies) have not been considered in such studies for COVID-19 prediction. Therefore, prediction for epidemic peak in most of studies has not been so accurate. In addition, these studies mainly predict the number of infectious persons from COVID-19 at particular region. However, the mortality (death rate) is also a crucial factor to take into account while forecasting the influence of a fatal pandemic i.e. COVID-19. In this paper, we propose an improvised five compartment mathematical model i.e. Susceptible (S)-Exposed (E)-Infected (I)-Recovered (R)-Death (D) (SEIRD model) to analyze the progression of COVID-19 and forecast the epidemic peak of COVID-19 pandemic under the influence of nationwide lockdown in India.

## MATERIALS AND METHODS

### Data collection

The data has been collected from the open source COVID-19 dataset official websites [4-6, 23-25]. Global data of COVID-19 has been retrieved from World Health Organization (WHO) official dashboard [5] whereas Ministry of Health and Family welfare (MOHFW) of India and other official data resources [25-27] has been used to study COVID-19 in India.

Figure (1) represents the spatial transmission of COVID-19 cases in India during three time periods i.e. pre-lockdown (January 30-March 24, 2020), lockdown (March 25-May 31, 2020) and post-lockdown period (June 1-July 10, 2020). It clearly indicates that COVID-19 cases were least transmitted during lockdown period as compared to pre-lockdown and post-lockdown across different states of India.

**Figure 1:**
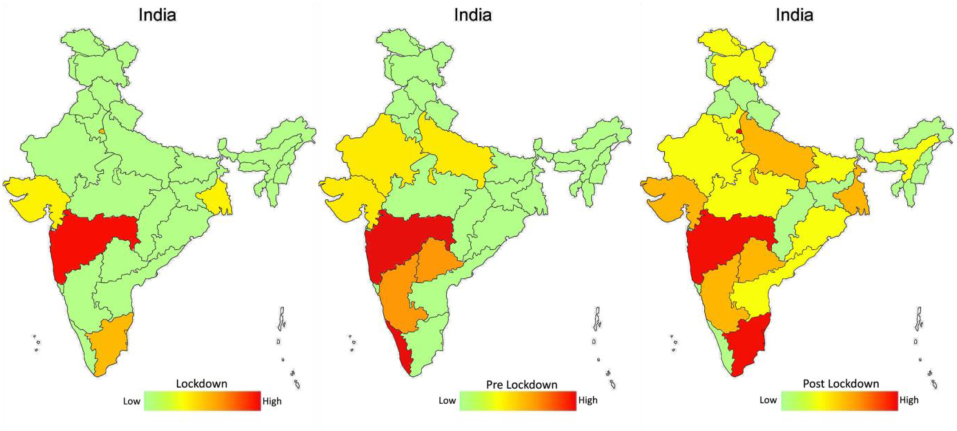
Spatial distribution of COVID-19 cases under the effect of lockdown in India.

### Mathematical model

We use five compartment epidemic model i.e. Susceptible (S)-Exposed (E)-Infected (I)-Recovered (R)-Death (D) (SEIRD model) to study COVID-19 in India. The SEIRD model has been improvised for accounting the effect of containment policy (lockdown) by adjusting the contact rate (*β*) and reproduction number (R) accordingly. Figure 2 represents the pictorial view of SEIRD model.

**Figure 2:**
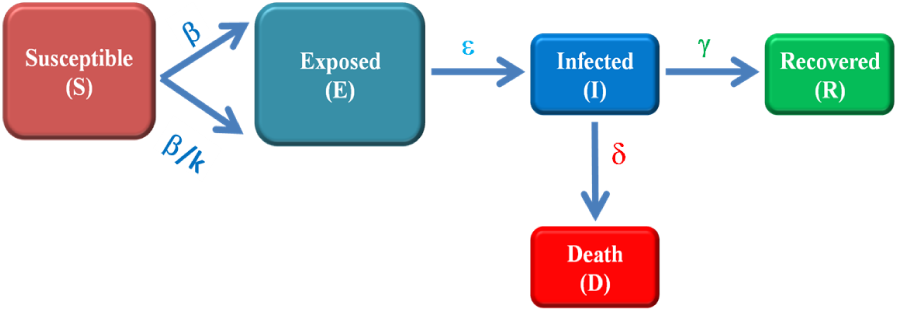
Schematic of modified SEIRD model.

Theoretically, SEIRD model can be represented by four differential equations [20]:

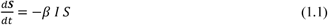

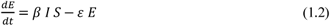

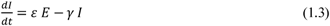

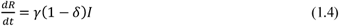

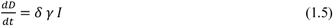

Here *β,ε,γ* are *δ* contact rate, incubation rate, recovery rate and death rate respectively and known as model parameters. Initial constraints are considered as *S(0) > 0, E(0) ≥0, I(0) ≥0, R(0) ≥ 0* and *D(0) ≥ 0*. Equations (1.1-1.5) are solved in Matlab software using function *“ode45”* and the contact rate *β* has been optimized using tool *“fminisearch”*. We have assumed that *β* has been reduced to, *β*/*k* (2≤ *k* ≤ 6) during lockdown period in India. The recovery rate (*γ*) and death rate (*δ*) has been calculated with the help of figure 3(a) and figure 3(b) respectively.

**Figure 3:**
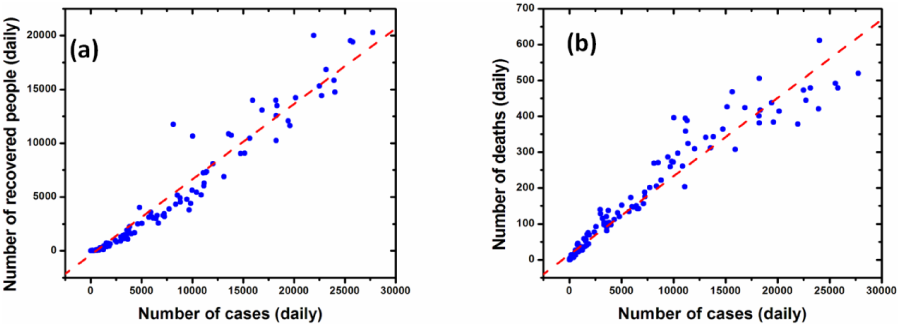
Determination of recovery rate and death rate in India (a) Reported recovered cases (b) Reported deaths with respect to daily confirmed cases.

The estimated or optimized model parameters are described in table (1) as:

**Table 1:** SEIRD model parameters used in equations (1.1-1.5)

| S. No. | SEIRD model parameters | Value |
| --- | --- | --- |
| 1 | $\beta$ (t) | 0.4809 |
| 2 | $\varepsilon$ | 0.1923 |
| 3 | $\gamma$ | 0.7012 |
| 4 | $\delta$ | 0.0215 |

### The time dependent basic reproduction number [R(t)]

The basic reproduction number (R) is most crucial parameter in SEIRD mathematical model. It determines how the disease is transmitting over population during a particular time interval. In our study, we have considered R as R(t) i.e. as a function of time. It is given as

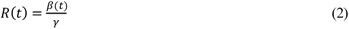

Case Fatality Risk/ratio (CFR) is another important parameter for epidemic study. It interprets the status of epidemic in terms of deaths. It is defined as

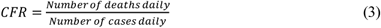

## RESULTS

At first, we have performed data based study of COVID-19 progression in India during the time period of January 30-July 10, 2020 i.e. 160 days. We have observed the trend of COVID-19 under the impact of lockdown. Figure 4 (a) represents the number of cases progression over time and indicates that the cases are very less during the lockdown period cases have raised abruptly just after lockdown has been lifted. Similarly, figure 4 (b) depicts the testing samples under lockdown effect. It has been observed that comparatively less samples have been tested during lockdown period, which indicates there might be underreported cases in India as well. However, Figure 4 (c) indicates that CFR is very low (1-3%) in India i.e. COVID-19 fatality is below average as compared to other countries in world.

**Figure 4:**
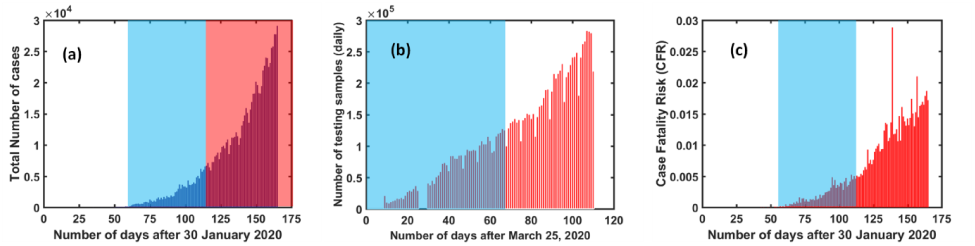
COVID-19 reported data [5] based analysis in during different time intervals in India (a) Reported number of daily cases (b) daily tested samples (c) CFR. [Blue shaded region represents actual lockdown period and red shaded region represents unlock period i.e. post lockdown period in India].

Further, the trend of time dependent reproduction number [R(t)] has been shown in figure 5(a). A descending trend has been observed in R during lockdown period whereas it has increased after the lockdown being lifted i.e. during post lockdown period. Further observation suggests that R(t) approaches unity around August 25, 2020. It indicates that the epidemic peak should be around August 2020 in India. Figure 5(b) represents the possible under-reported cases in India. It can be defined as the difference in the model predicted cases and reported cases. A significant number of underreported cases (∼10^5^) has been observed in beginning of post lockdown in India.

**Figure 5:**
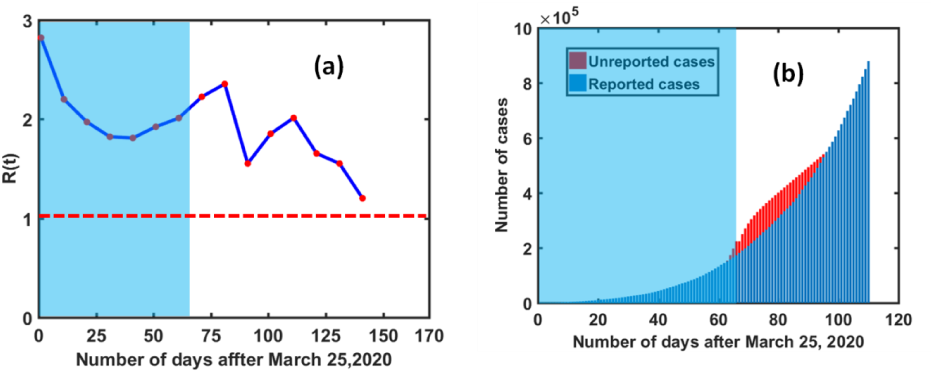
COVID-19 reported data based analysis in during different time intervals in India (a) Basic Reproduction number variation (b) underreported cases.[Blue shaded region represents actual lockdown period].

### Prediction of COVID-19 epidemic peak in India

To forecast the trajectory of COVID-19 in subsequent months, we have applied SEIRD model to predict the trajectory of COVID-19 under the nationwide lockdown in India. Figure 6(a) represents the actual estimated trajectory of all five compartments (susceptible, exposed, infected, recovered and death) COVID-19 in India Figure 6(b) provides more clear insight of epidemic peak in India based on actual parameters. It has been estimated that maximum number of COVID-19 cases will be found around 2.5 ×10^6^ at mid-August 2020. The red colored markers represent the COVID −19 trajectory based on reported data [6].

**Figure 6:**
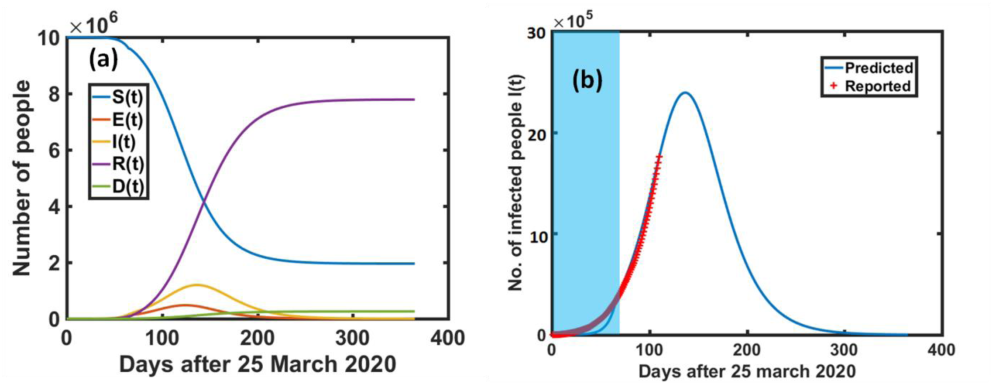
Mathematical model prediction of COVID-19 dissemenation in India (a) Five compartment trajectory over time (b) COVID-19 epidemic peak prediction in India.[Blue shaded region represents actual lockdown period].

Figure 7 (a) shows the five compartment (SEIRD) trajectory under no lockdown condition in India. It indicates that the number of cases would be around 10^7^ in that case. Moreover, if lockdown of 100 days could have been implemented instead of 65 days, the number of cases would be very less i.e. 10^5^. However, the peak would have been shifted further (figure 7(b)).

**Figure 7:**
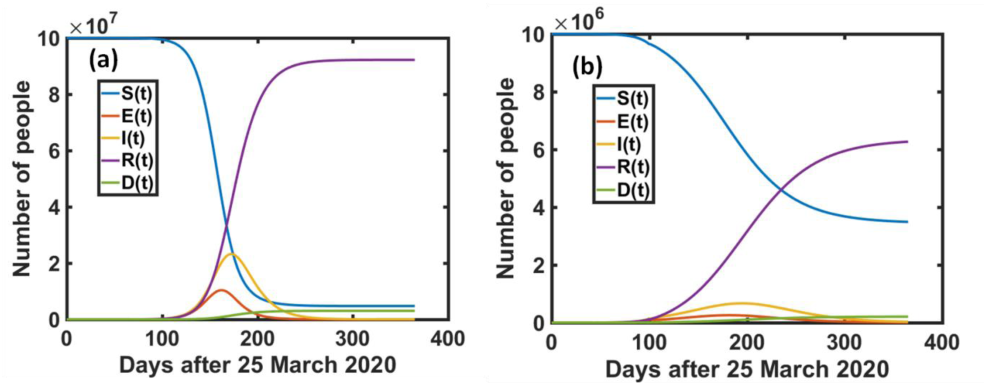
Mathematical model prediction of COVID-19 dissemenation in India under (a) without lockdown (b) Lockdown of 100 days.

Table 2 represents the predicted numerical data under various lockdown possiblities in India.Observations indicate that the cases could be order of 10^7^, if lockdown has not been implemented in India whereas lockdown of 100 days instead of 65 days could suppress the cases up to 50% of actual cases prediction in India. It confirms that the number of positive cases can be controlled in a better manner under lockdown period.

**Table 2:** Epidemic peak prediction using SEIRD model in India under lockdown effect

| S.No. | Lockdown period | Estimated COVID-19 epidemic peak |  |
| --- | --- | --- | --- |
|  |  | Date | Estimated maximum cases |
| 1 | 0 days (hypothetical cases i.e. if no lockdown has been implemented in India) | September 25, 2020 | $2.1 \times 10^7$ |
| 2 | 65 days (March 25- May 31 2020) (actual case) | August 16, 2020 | $2.5 \times 10^6$ |
| 3 | Lockdown of 100 days (extensive case) | September 15, 2020 | $1.2 \times 10^6$ |

## CONCLUSION AND DISCUSSION

In this study, we have analyzed and predicted COVID-19 dissemination in India using a five compartment mathematical model i.e. SEIRD model. The conventional SEIRD model has been improvised for accounting the impact of COVID-19 mitigation policy i.e. nationwide lockdown in India. The modified transmission rate [β(t)] and time dependent reproduction number [R (t)] are the key elements of our improvised SEIRD model. Moreover, the data has been examined up to July 10, 2020 in this study. The obtained results are in good agreement with the reported data. Moreover, COVID-19 trajectory has also been estimated under different possible lockdown scenario. Corresponding outcomes indicate that the cases could have been controlled up to a better extent with extensive lockdown period. Meanwhile nationwide lockdown has been implemented at emerging phase of COVID-19 in India, but only for sixty five days. Keeping in mind the population and medical facilities in India, such short lockdown period might not be sufficient. Our study suggests that the lockdown could have been extended up to hundred days. In short, our study confirms that nationwide lockdown is an effective controlling policy for COVID-19 in India and will definitely plays a crucial role in epidemic peak suppression of COVID-19 dissemination in India. Although our improvised SEIRD model predicts satisfactory results but one can include few additional parameters i.e. imported or exported cases, under-reported cases etc. to enhance the validity of mathematical model while predicting an epidemic dissemination.

## Data Availability

Publicly available datasets were analyzed in this study. This data can be found here: https://data.world/kim4597/world-covid19/workspace/intro.

## AUTHOR CONTRIBUTIONS

VT, ND, NS designed the study. VT, NS developed the mathematical model and simulation coding. ND has contributed in manuscript writing and data interpretation.

